# Development and potential usefulness of the COVID-19 Ag Respi-Strip® diagnostic assay in a pandemic context

**DOI:** 10.1101/2020.04.24.20077776

**Authors:** Pascal Mertens, Nathalie De Vos, Delphine Martiny, Christian Jassoy, Ali Mirazimi, Lize Cuypers, Sigi Van Den Wijngaert, Vanessa Monteil, Pierrette Melin, Karolien Stoffels, Nicolas Yin, Davide Mileto, Sabrina Delaunoy, Henri Magein, Katrien Lagrou, Justine Bouzet, Gabriela Serrano, Magali Wautier, Thierry Leclipteux, Marc Van Ranst, Olivier Vandenberg, LHUB-ULB SARS-COV-2 working diagnostic group, Béatrice Gulbis, Françoise Brancart, François Bry, Brigitte Cantinieaux, Francis Corazza, Fréderic Cotton, Maud Dresselhuis, Bhavna Mahadeb, Olivier Roels, Jacques Vanderlinden

## Abstract

**Introduction:** COVID-19 Ag Respi-Strip, an immunochromatographic (ICT) assay for the rapid detection of SARS-CoV-2 antigen on nasopharyngeal specimen, has been developed to identify positive COVID-19 patients allowing prompt clinical and quarantine decisions. In this Original Research article, we describe the conception, the analytical and clinical performances as well as the risk management of implementing the COVID-19 Ag Respi-Strip in a diagnostic decision algorithm.

**Materials and Methods:** Development of the COVID-19 Ag Respi-Strip resulted in a ready-to-use ICT assay based on a membrane technology with colloidal gold nanoparticles using monoclonal antibodies directed against the SARS-CoV and SARS-CoV-2 highly conserved nucleoprotein antigen. Four hundred observations were recorded for the analytical performance study and thirty tests were analysed for the cross-reactivity study. The clinical performance study was performed in a retrospective multi-centric evaluation on aliquots of 328 nasopharyngeal samples. COVID-19 Ag Respi-Strip results were compared with qRT-PCR as golden standard for COVID-19 diagnostics.

**Results:** In the analytical performance study, the reproducibility showed a between-observer disagreement of 1.7%, a robustness of 98%, an overall satisfying user friendliness and no cross-reactivity with other virus-infected nasopharyngeal samples. In the clinical performance study performed in three different clinical laboratories we found an overall sensitivity and specificity of 57.6% and 99.5% respectively with an accuracy of 82.6%. The cut-off of the assay was found at Ct<22. User-friendliness analysis and risk management assessment through Ishikawa diagram demonstrate that COVID-19 Ag Respi-Strip may be implemented in clinical laboratories according to biosafety recommendations.

**Conclusion:** The COVID-19 Ag Respi-Strip represents a promising rapid SARS-CoV-2 antigen assay for the first-line diagnosis of COVID-19 in 15 minutes. Its role in the proposed diagnostic algorithm is complementary to the currently-used molecular techniques.

## INTRODUCTION

Severe acute respiratory syndrome coronavirus 2 (SARS-CoV-2) represents a major health threat for humankind (1). In the absence of vaccine and specific antiviral treatment, the containment of the pandemic relies mainly on rapid identification and isolation of COVID-19 patients (2). In addition to chest computed tomography (CT), this strategy is based on the availability of real-time reverse transcription polymerase chain reaction (qRT-PCR) to be performed on any suspect patient presenting specific symptoms (3). These symptoms being similar to seasonal flu, it’s currently not possible to test all patients with flu-like symptoms due to the lack of resources and available diagnostic tests. As mentioned in the audio interview of the New England Journal of Medicine the 19^th^ of March 2020 (2), the importance of establishing the correct diagnosis is central to give the appropriate care to COVID-19 patients.

So far, several molecular-based tests have been developed and are being implemented in laboratories and reference centres with capabilities to perform such tests (see https://www.who.int/emergencies/diseases/novel-coronavirus-2019/technical-guidance/laboratory-guidance for details). However, the availability of molecular diagnostic tests is a concern, as we face worldwide shortage of the reagents. Although molecular diagnosis is the most sensitive and specific diagnostic method, the need for material, reagent and trained personnel limit the number of assays that can be performed and saturates the laboratories. Moreover, qRT-PCR still does not have a very rapid turnaround time (TAT).

The development of rapid diagnostic assays allows faster confirmation of a clinical suspicion of COVID-19, leading to earlier isolation and appropriate clinical care for the patients with positive results. Several serological tests have been developed but serological antibody-detection assays do not fulfil the requirement of the detection early after infection as the average incubation period of 3-5 days is too short for the development of an immune response (4).

In this perspective, Coris BioConcept (a Belgian manufacturer) has developed an immunochromatographic assay (ICT) for the rapid detection of SARS-CoV-2 antigen on nasopharyngeal specimens in about 15 minutes. Thanks to the results from previous research work on SARS-CoV, the nucleoprotein was identified as the best target for a sensitive diagnostic sandwich assay using monoclonal antibodies (5, 6,7). The SARS-CoV-2 shares high similarity with bat coronaviruses and the known SARS-CoV of the 2002-2003 epidemic (8) provided the opportunity to use previously developed reagents for developing a rapid diagnostic assay able to also detect the new SARS-CoV-2. The diagnostic technique, consisting of an anti-SARS-CoV capture antibody fixed onto nitrocellulose strip and a labelled anti-SARS-CoV antibody migrating with the buffer and the sample.

Regarding the COVID-19 pandemic and the urgency of sharing relevant data, in this original research article we describe the analytical performance of COVID-19 Ag Respi-Strip, according to the requirements of the actual European Directive 98/79/EC (9), the future European Regulation 2017/746 on *in vitro* diagnostic (IVD) medical devices (10), the Scandinavian SKUP-protocol (11) used for validation of qualitative tests and the clinical performance obtained with a multi-centric retrospective study. In addition, we reflect on the risk management and the conditions to be fulfilled before implementation as a point-of-care test (POCT) outside the hospital.

## MATERIALS AND METHODS

### Development of the COVID-19 Ag Respi-Strip

#### Antibodies and antigen

Eleven antibodies (designed A to K) (12) were coated at various concentrations on nitrocellulose (Advanced microdevices, India) with antibodies A to J coupled to colloidal gold beads (NanoQ, Belgium). Recombinant SARS-CoV nucleoprotein (recNP) preparation was obtained as described previously (12) and was coated on a nitrocellulose membrane or conjugated on colloidal gold nanoparticles. Recombinant his-tagged SARS-CoV-2 NP (recNP-2) has been produced in insect cell and purified (2-step purification); final purity > 90% (Genscript, Leiden, NL).

#### COVID-19 Ag Respi-Strip

the ICT strip consists of nitrocellulose laminated on a plastic backing, colloidal-gold conjugates being dried in a conjugate pad (Ahlstrom-Munksjö, France) overlapping the bottom of the nitrocellulose. For preliminary direct detection, SARS-CoV and SARS-CoV-2 NPs were coated at 100µg/ml and gold-labelled antibodies were deposited at 0.85µl/mm at 3 OD530nm. For the COVID-19 Ag Respi-Strip test, monoclonal antibodies directed against SARS-CoV and SARS-CoV-2 highly conserved nucleoprotein antigen are coated on the nitrocellulose. Another monoclonal antibody is conjugated to colloidal gold nanoparticles. The conjugate is immobilized on the conjugate pad. During the development, tests analysing the antibody reactivity and intensity were performed using serial dilutions of SARS-CoV-2 in a final volume of 300µl of buffer (data not shown here, cfr. Supplementary material). The results were determined after 15 min.

The standard operating procedure for COVID-19 Ag Respi-Strip is as follows (Figure 1): Transfer 100 µL of nasopharyngeal sample (nasopharyngeal aspirates, nasopharyngeal washes or nasal/nasopharyngeal swabs) in the collection tube. Add 100 µL of the LY-S dilution buffer to reach a dilution ratio of 1/2. Cap the tube with the stopper. Stir thoroughly to homogenize the solution. Open the tube. Immerse the strip in the direction indicated and close the tube with the stopper. Allow to react 15 minutes and read the result. For the interpretation of results, with a negative test result, a reddish-purple line appears at the Control line (C) position (upper line). No other band is present. For a positive test result, in addition to a reddish-purple band at the Control line (C), a visible reddish-purple band appears at the Test line position (T). Intensity of the test line may vary according to the quantity of antigens found in the sample. Any reddish purple line (T), even weak, should be considered as a positive result. An invalid test result is when the absence of a Control line indicates a failure in the test procedure. Repeat invalid tests with a new strip. Discard the closed tube according to Biohazard rules.

**Figure 1:**
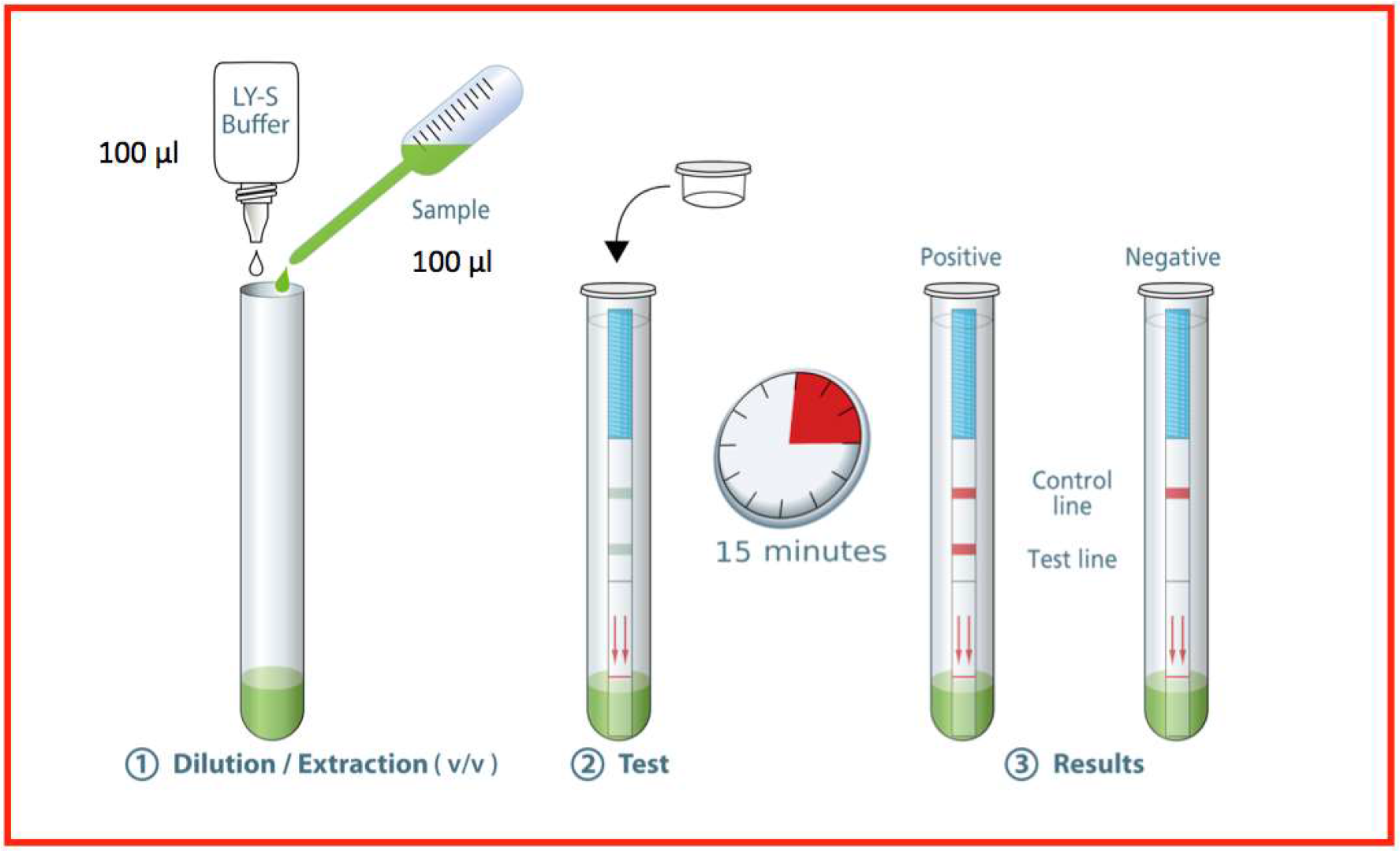
Standard Operating Procedure SARS-CoV-2 Respi-Strip from Coris BioConcept.

#### ELISA

A 96-well microtiter plate was coated with 50 µL recNP and recombinant maltose binding protein (MBP, 1 µg/mL) and incubated overnight at 4 °C. The plate was washed with water and washing buffer (phosphate-buffered saline/0.5% Tween-20, PBS-T) and 200 µL blocking solution (PBS-T/5 % milk powder) was added for 20 minutes. Blocking solution was discarded and mAbs were added at 100 pg/ml in 50 µL blocking solution to recNP and MBP and incubated for 1 hour. The plate was washed again and 50 µL rabbit anti-mouse IgG/HRP (Dako) was added at 1:1000 dilution for another 1 hour incubation. The plate was washed and 50 microliter TMB substrate solution was added. All incubations took place at room temperature. The enzymatic reaction was stopped by addition of 50 µL 1 N H2SO4 and light absorption was measured with a photometer at 450 nm using 570 nm as reference wavelength. Measurements were done in duplicates. To obtain the final OD, the OD obtained with the control protein MBP was subtracted from the OD obtained with recNP.

#### Virus

SARS-CoV-2 passage 3 (SARS-CoV-2-Iso_01-Human-2020-02-07-Swe, accession no/genebank no MT093571) was cultured on Vero E6 cells. The titre was determined using plaque assay as describe before with fixation of cells 72hpi. All experiments involving isolated SARS-CoV-2 were performed in Biosafety Level 3 Laboratory at Public Health Agency of Sweden (Folkhälsomyndigheten, Stockholm, Sweden).

#### qRT-PCR

Sample was extracted using Direct-zol RNA MiniPrep kit (Zymo Research). qRT-PCR was run using E-gene SARS-CoV-2 primers/probe following World Health organization advises (13).

### Analytical performance study

During the development phase, analytical performance was first performed on serial dilution of cultured virus in parallel of tittering the same virus preparation on Vero E6 cells and testing by qRT-PCR.

For the analytical sensitivity and specificity obtained in a clinical biology lab setting, 60 samples from UZ Leuven, the National Reference Centre for the diagnosis of COVID-19 in Belgium were analysed in the laboratory LHUB-ULB (Laboratoire Hospitalier Universitaire Bruxelles - Universitair Laboratorium Brussel), Brussels. All samples were nasopharyngeal swabs in viral transport medium. The analysis protocol with the COVID-19 Ag Respi-Strip was as follows: of the 20 positive patient samples with Cycle threshold (Ct) below 25, ten of them were analysed in duplicate; of the 20 weakly positive samples with 25<Ct<37.7 all were analysed in duplicate; and of the 20 negative patient samples, ten of them were analysed in duplicate. Duplicate specimen were randomly chosen. For these 100 analyses, 4 observers delivered a qualitative result, resulting in 400 observations. The diagnostic efficacy of the COVID-19 Ag Respi-Strip was evaluated comparing the results with those previously obtained on fresh nasopharyngeal samples tested for SARS-COV-2 with the reference qRT-PCR test (13, slightly adapted in NRC). Regarding the qRT-PCR reference test, total nucleic acid was extracted using NucliSens extraction on easyMAG (BioMérieux, Lyon, France), followed by addition of a Phocine Distemper Virus (PDV) internal control (IC) (14). PCR amplification was performed on QuantStudio Dx (Thermo Fisher Scientific), using slightly adapted E-gene primers and probe (13). After the run, amplification plots were analyzed and interpreted using QuantStudio Test Development Software (version 1.0, Thermo Fisher Scientific).

ICT assay were performed following the manufacturer’s instructions using high containment measures (Biological Safety Cabinet Class II). With these samples, the analytical performance study consisted of analytical sensitivity, analytical specificity, reproducibility (between-observer disagreement with 4 observers simultaneously reading the result) and robustness.

For the cross-reactivity study, experiments to assess the reactivity of COVID-19 Ag Respi-Strip to other pathogens were conducted in the LHUB-ULB depending on specimen availability (N=30, consisting of 20 nasopharyngeal aspirates and 10 sputa). Clinical residual anonymized respiratory samples from patients with non-SARS-COV-2 infections were tested. The concentrations of the pathogens fluctuated owing to the available stock, and the clinical specimen with the highest virus load were selected.

The User friendliness study was performed according to the Scandinavian protocol SKUP/2004/35* with 5 questioned operators responding independently to a checklist (11).

### Clinical performance study

In order to consider the implementation of the COVID-19 Ag Respi-Strip into the national diagnostic algorithm, an urgent multi-centric retrospective study aiming to assess the clinical performance of this rapid assay against current molecular methods (golden standard) was performed. Overall, 328 nasopharyngeal samples from patients suspected of SARS-COV-2 infections attending from 19 to 30 March 2020 in three university laboratories located in Belgium were tested following manufacturer’s instruction to assess the clinical sensitivity, clinical specificity, positive predictive value (PPV), negative predictive value (NPV) and accuracy in order to propose a diagnostic algorithm adapted at the current situation. This retrospective multi-centric evaluation integrates 322 randomly selected nasopharyngeal swabs (NPS) (flocked swab + UTM 3 mL (or 1mL of Amies) (Copan, Brescia, Italy)), 4 nasopharyngeal aspirate (NPA) (diluted with 3 mL of viral transport medium composed of veal infusion broth (Difco, Becton Dickinson, Sparks, MD, USA) supplemented with bovine albumin (Sigma Aldrich, St Louis, MO, USA)) (15) and 2 Broncho-Alveolar Lavage (BAL) of the biobanks at LHUB-ULB, UZ Leuven and CHU Liège. Aliquots of these patient samples were analysed with COVID-19 Ag Respi-Strip and compared to the qRT-PCR result.

At the laboratory LHUB-ULB, viral RNA extraction was performed by QIAsymphony DSP Virus/Pathogen Kit (QIAGEN) which extracted 400µl sample eluted in 60µl elution buffer for the Mini Kit and 800µl sample eluted in 110µl buffer for the Midi Kit, or by m2000 mSample Preparation SystemDNA Kit (Abbott) using 1000µl manually lysed sample (700µl sample + 800µl lysis buffer from kit) eluted in 90µl elution buffer. A qRT-PCR internal control was added at each extraction. qRT-PCR was performed using 10µl of extracted sample in the RealStar® SARS-CoV-2 RT-PCR Kit from altona-diagnostics with a cut-off set at 40 Ct.

The RT-PCR protocol used in Liege for the comparison was as follow: RNA was extracted from clinical samples (300µL) on a Maxwell 48 device using the Maxwell RSC Viral TNA kit (Promega). Reverse transcription and RT-PCR were performed on a LC480 thermocycler (Roche) based on Charité’s protocol for the detection of RdRp and E genes (16) using the Taqman Fast Virus 1-Step Master Mix (Thermo Fisher).

For UZ Leuven, a second qRT-PCR method was performed on Panther Fusion (PF, Hologic, San Diego, USA) Open AccessTM SARS-CoV analysis. Analytical protocol was as follow: 500μL UTM from the nasopharyngeal sample is added to a PF lysis tube, mixed by pipetting and loaded on the instrument. All following steps, including total nucleic acid extraction, reverse transcription and real-time PCR, are automatized on the instrument and were defined in the LDT-protocol using the myAccess software.

This SARS-CoV assay targets 2 SARS-CoV genes: the E-gene for which primers and probers are slightly adapted from Corman et al. (13), and the gene ORF1-b. Amplification plots are analysed by the system software using parameters defined in the LDT-protocol. A linear regression line y = 0.9993x + 5.4341 was constructed to normalize the difference in Ct values found with the two methods used in UZ Leuven (y=Panther Fusion and x=Quantstudio). To obtain a cut-off, all positive qRT-PCR results were grouped per category of one Ct and matched with the most frequent qualitative interpretation obtained with COVID-19 Ag Respi-Strip. At least 50% of qualitative interpretations are found positive at the cut-off.

The study was approved by the ethical boards (P2020/191 for Hôpital Erasme, CE2020/65 for CHU Brugmann, AK/10-06-41/3907 for CHU Saint-Pierre, S63896 for UZ Leuven. For CHU of Liege, no specific approval was requested to the EC as a leaflet including the following statement is given to all admitted patients: “*According to the law of the 19th December 2008, any left-over of biological material collected from patients for their standard medical management and normally destroyed when all diagnostic analysis have been performed, can be used for validation of methods. The law authorizes such use except if the patient expressed an opposition when still alive (presume consent)”*.

### Risk management

Risks and bottlenecks of the use of COVID-19 Ag Respi-Strip will be presented in an Ishikawa diagram (17).

## RESULTS

### Selection and Characterization of Monoclonal Antibodies

Four different assays were performed to assess the reactivity of antibodies towards SARS-CoV NP (I-III) and SARS-CoV-2 (IV) nucleoproteins. Antibodies were tested using an ELISA on immobilized recNP showing various reactivitiess (Supplementary Digital Content: Table 1, column I). Prior to the use of antibodies in a sandwich detection assay, antibodies were individually tested for their ability to capture and/or detect the target recNP in ICT format. Antibodies were thus coated onto nitrocellulose and tested using the recNP coupled to colloidal gold beads; reactivity was recorded as based on visual intensity on the ICT strips (Suppl. Table 1, column II). Lastly, antibodies were coupled to colloidal gold beads and migrated on an ICT strip where the recNP was immobilized; results are recorded in Suppl. Table 1, Column III. As can be observed, the reactivity of antibodies in the ELISA assay does not predict their ability to work properly as capture or detection reagent in an ICT format as described previously for other targets (12). Moreover, antibodies with no detectable activity as detection reagent (antibodies B, C, D, column III) show weak to good capture capability (Column II). Comparing reactivity on both recNP and recNP-2, the overall reactivity was higher on recNP-2 although antibodies were initially elicited and selected against recNP. This may reflect different protein preparation protocols. More interestingly, antibodies B, C and H that were reacting against recNP did not at all react with recNP-2 leading to the hypothesis that these antibodies may react with an epitope which is specific for recNP and not present on recNP-2.

**Table 1.** Analytical performance study

|  | UZ Leuven specimen |  | LHUB-ULB test |  | New COVID-19<br>Ag Respi-Strip<br>(Saline Buffer) |
| --- | --- | --- | --- | --- | --- |
|  | Number of<br>samples | Lab<br>Interpretation<br>(qRT-PCR) | Observers | N<br>observations | Results |
| **Positive samples**<br>**(with qRT-PCR)** |  |  |  |  |  |
| CT< 25 | 30 | Positive | 4 | 120 | 89/120 (74.2%) |
| 25<CT<37.7 | 40 | Weak positive | 4 | 160 | 20/160 (12.5%) |
| **Negative samples** | 30 | Negative | 4 | 120 | 120/120 (100.0%) |

Sandwich ICT assays were performed by combining antibodies working as capture reagents (coated on nitrocellulose) and antibodies working as detection reagent (gold-labelled). These ICT were tested using recNP at a similar concentration for all tests (100ng/ml). All 20 combinations giving a visible signal were assessed on their ability to detect SARS-CoV-2 virus (Suppl. Table 2).

**Table 2.**
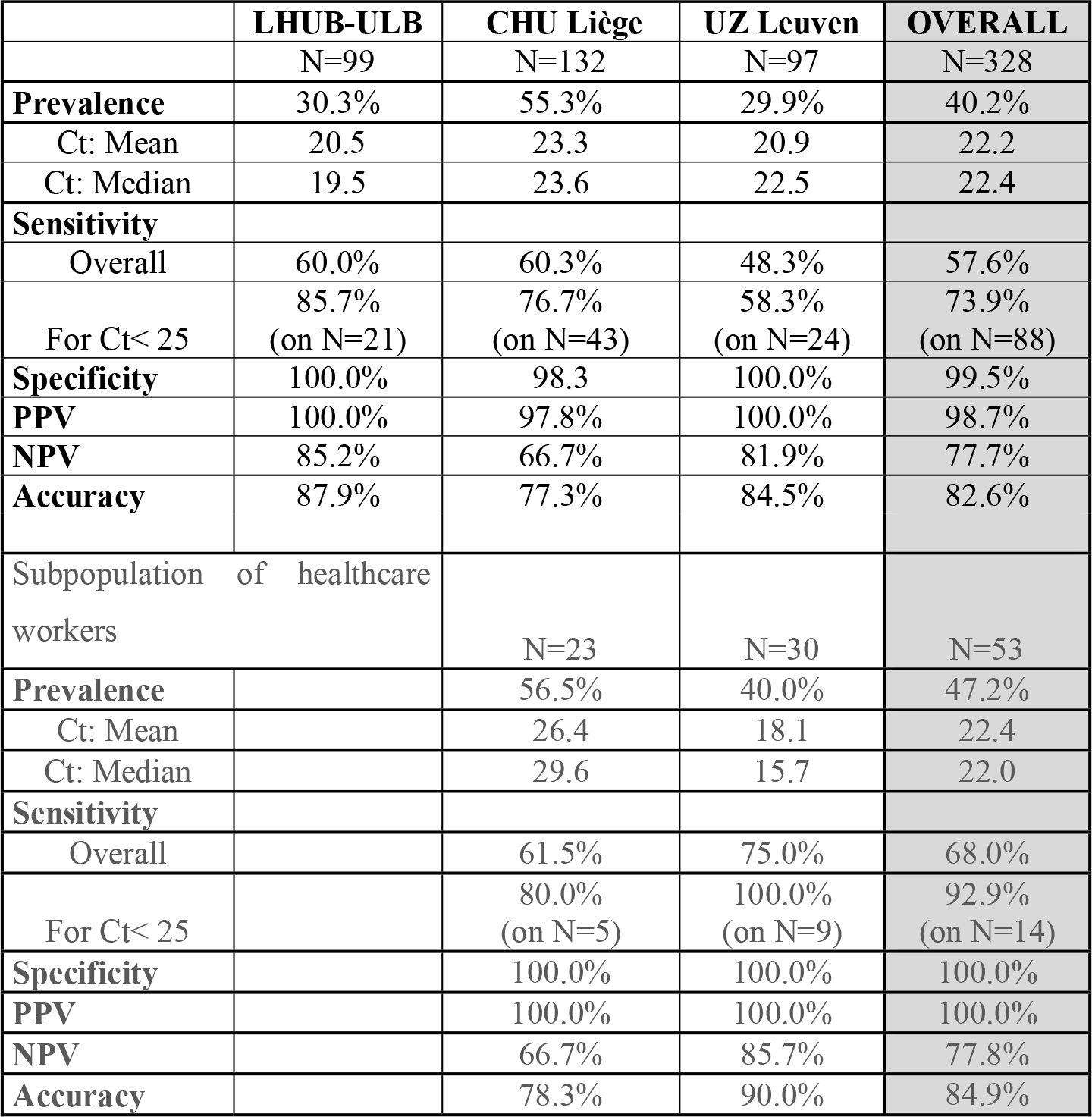
Clinical performance study

|  | **LHUB-ULB** | **CHU Liège** | **UZ Leuven** | **OVERALL** |
| --- | --- | --- | --- | --- |
|  | N=99 | N=132 | N=97 | N=328 |
| **Prevalence** | 30.3% | 55.3% | 29.9% | 40.2% |
| Ct: Mean | 20.5 | 23.3 | 20.9 | 22.2 |
| Ct: Median | 19.5 | 23.6 | 22.5 | 22.4 |
| **Sensitivity** |  |  |  |  |
| Overall | 60.0% | 60.3% | 48.3% | 57.6% |
| For Ct< 25 | 85.7%<br>(on N=21) | 76.7%<br>(on N=43) | 58.3%<br>(on N=24) | 73.9%<br>(on N=88) |
| **Specificity** | 100.0% | 98.3 | 100.0% | 99.5% |
| **PPV** | 100.0% | 97.8% | 100.0% | 98.7% |
| **NPV** | 85.2% | 66.7% | 81.9% | 77.7% |
| **Accuracy** | 87.9% | 77.3% | 84.5% | 82.6% |
| Subpopulation of healthcare workers |  | N=23 | N=30 | N=53 |
| **Prevalence** |  | 56.5% | 40.0% | 47.2% |
| Ct: Mean |  | 26.4 | 18.1 | 22.4 |
| Ct: Median |  | 29.6 | 15.7 | 22.0 |
| **Sensitivity** |  |  |  |  |
| Overall |  | 61.5% | 75.0% | 68.0% |
| For Ct< 25 |  | 80.0%<br>(on N=5) | 100.0%<br>(on N=9) | 92.9%<br>(on N=14) |
| **Specificity** |  | 100.0% | 100.0% | 100.0% |
| **PPV** |  | 100.0% | 100.0% | 100.0% |
| **NPV** |  | 66.7% | 85.7% | 77.8% |
| **Accuracy** |  | 78.3% | 90.0% | 84.9% |

Assay on virus was performed using a serial dilution of culture supernatant. The relative intensities observed on recNP and on SARS-CoV-2 were similar, the prototypes 4 and 5 giving the highest signals. COVID-19 Ag Respi-Strip, corresponding to the Prototype 5, was further characterised for analytical performance during development of the IVD medical device.

### Analytical performance study

During the development phase, the analytical performance was first performed on two-fold serial dilutions of recNP-2. The limit of detection was defined as the last dilution tested positive (15 tests, 2 independent readers). The assay was shown to have a detection level down to 250 pg/ml. The assay was tested on supernatant of Vero E6 cells infected with SARS-CoV-2 virus. We have found that the assay can detect 5 ×10e3 pfu/mL corresponding to a Ct value of 23.7 in a qRT-PCR assay.

During validation in the clinical biology lab, all the 130 strips used in this analytical study were valid, except for two. Analytical sensitivity for patients having a high viral load (Ct<25) was 74.2% with an analytical specificity of 100.0% (Table 1). The reproducibility showed a between-observer disagreement of 1.7 % (N= 7/398). The robustness is 98.0% showing only two out of 100 tests being invalid, as they lacked the visual test control line because migration did not succeed In all of the succeeded tests (N=98/98), the reaction fulfilled in time, i.e. after 15 minutes.

No cross-reactivity nor interference has been found in nasopharyngeal samples containing the following pathogens (overall N=20): Coronavirus HKU1 (N=2), Adenovirus (N=3), Enterovirus (N=2), Influenza A virus (N=3), Influenza B virus (N=3), human Metapneumovirus (N=2), Parainfluenza virus (N=1), Rhinovirus (N=4), RSV (N=2), and *Mycoplasma pneumoniae* (N=1).

Because of the unavailability of nasopharyngeal samples with *Staphylococcus aureus*, the cross-reactivity study couldn’t be properly performed, and the test circumstances were approximately simulated with N=10 sputa with abundant culture of *S. aureus*. The cross-reactivity for *S. aureus* in sputum, which isn’t the prescribed sample type for COVID-19 diagnostics and moreover has unknown matrix interference, gave one false positive test result. On the contrary, in all weakly and strongly positive SARS-CoV-2 observations with the nasopharyngeal samples of UZ Leuven, no false positive result has been detected with the COVID-19 Ag Respi-Strip (N=0/108).

Cross-reactivity was also checked for viruses from culture supernatants: Coronaviruses OC43, NL63, 229E and HKU1, and SARS-CoV. No cross-reactivity was observed for the four seasonal coronaviruses; cross-reactivity was observed for SARS-CoV as expected.

The user friendliness is satisfactory for the information in the instruction for use (IFU) of the manufacturer and for the time factors related to the pre-analytical and analytical phase. As the internal and external quality controls unrelated to the kit aren’t available yet, the overall rating for quality control is less satisfactory, although the interpretation of the quality control line integrated in the strip is satisfactory even if some control lines showed a weak intensity. The user friendliness evaluation related to the operation showed a very satisfactory rating (Supplementary digital content: Table 2).

### Clinical performance study

The overall clinical sensitivity is 57.6% and clinical specificity is 99.5%, with a PPV of 98.7%, NPV of 77.7% and an accuracy of 82.6% (Table 2). Even if the overall sensitivity of 57.6% will detect 6 out of 10 random people with COVID-like symptoms presenting at the hospital, in the subpopulation with the most contagious patients with the highest viral load (i.e. with Ct<25), this test will detect 7 out of 10 positive COVID-19 patients. In the COVID-19 population, the Cts ranged from 9.4 to 39.4 and the cut-off of the COVID-19 Ag Respi-Strip was found at Ct<22. Below the cut-off, the clinical sensitivity was 95.0%. A small number of samples has been studied from symptomatic healthcare workers (N=53), and the clinical performance showed a sensitivity of 68.0% and specificity of 100.0% in this subpopulation. Based on the results of the clinical performance study, a diagnostic decision algorithm is proposed in figure 2.

**Figure 2:**
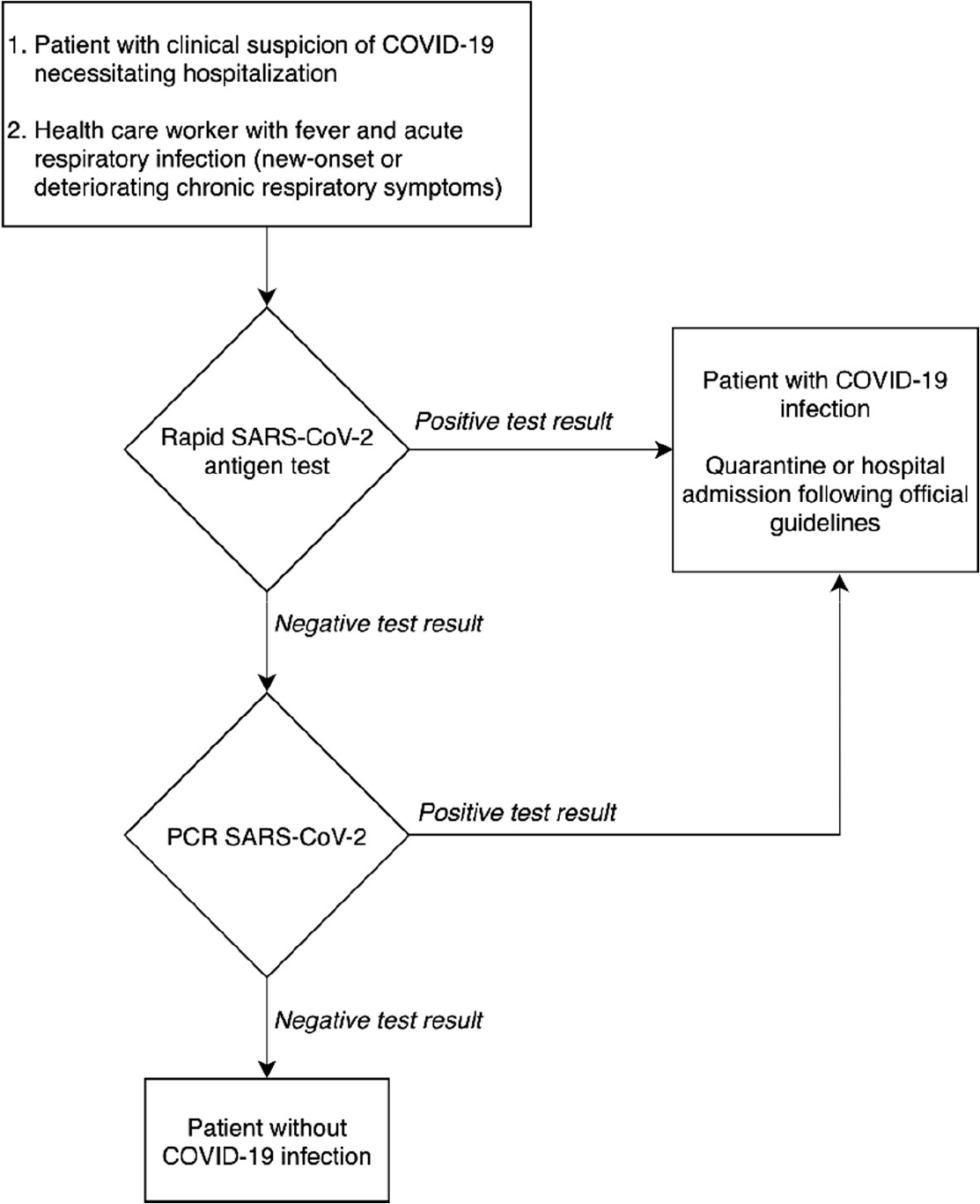
Proposal for a diagnostic decision algorithm.

### Risk Management

To visualize the risks and bottlenecks of the use of COVID-19 Ag Respi-Strip, an Ishikawa diagram has been created considering continuous risk management (Fig. 3). The facilities of a clinical biology laboratory are recommended in the Ishikawa diagram, to respond to the highest quality assurance and to permit lab technicians to handle the sample according to biosafety recommendations.

**Figure 3:**
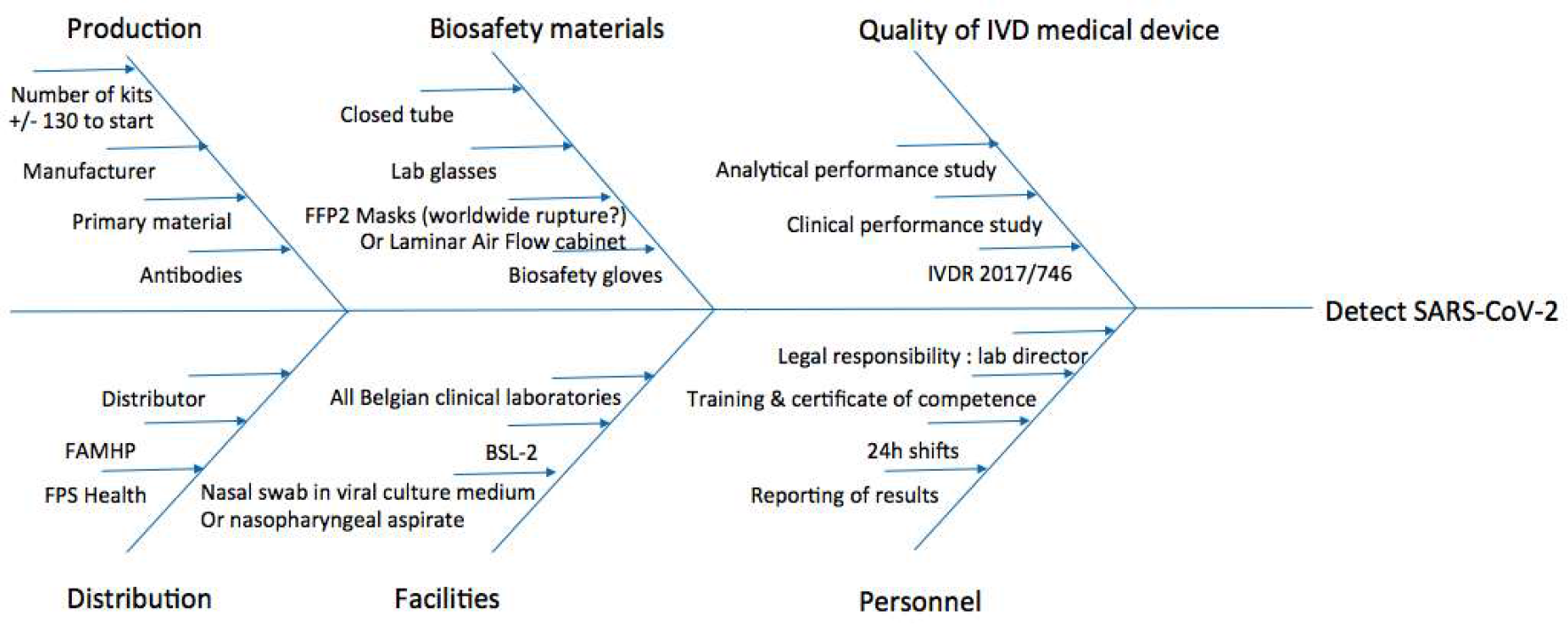
Ishikawa diagram of rapid SARS-CoV-2 diagnostic tests for Clinical Labs’ Implementation.

## DISCUSSION

The extraordinary spread of SARS-CoV-2 has resulted in the need for accelerated development of rapid and accurate laboratory diagnostic tests, allowing fast and accurate detection of infected patients (18). In this perspective, fast near-patient testing (POCT) could represent an effective diagnostic tool for prompt clinical and quarantine decisions. Besides the recent certification of molecular POCT such as Xpert^®^Xpress SARS-CoV-2 on GeneXxpert (Cepheid), or ID NOW COVID-19 on Alere-i (Abbott), antigenic test could represent a valuable alternative, especially in the context of worldwide reagents’ and instruments’ shortage (which have already been reported for these molecular POCTs).

Next to a pre-peer reviewed article reporting a fluorescence immunochromatographic assay detecting nucleocapsid protein of SARS-CoV-2 in nasopharyngeal swab (19), this original research report is, according to our knowledge, the first one describing the analytical and clinical performance of a non-fluorescent ICT that can detect SARS-CoV-2 antigen in nasopharyngeal samples.

The analytical sensitivity of 74.2% found in a subpopulation with high SARS-CoV-2 viral load (Ct<25), the analytical specificity of 100.0% and robustness of 98.0% for COVID-19 Ag Respi-Strip show a promising new technique. In addition, our multi-centric retrospective study confirms the promising results, with a clinical sensitivity of 57.6% and a clinical specificity of 99.5%. The clinical sensitivity is 68.0% in a sub-population of healthcare workers and increases to 73.9% in a subpopulation of the most contagious patients with high viral load (Ct<25). It should be noted that for the clinical validation in this study, the samples were collected in transport media first for being processed by qRT-PCR, and leftovers were used with some delay with COVID-19 Ag Respi-Strip. Not only the sensitivity may have been impacted by the delay between sample collections and processing, also the dilution of sample in the transport media may affect the sensitivity of the assay. A clinical validation using the swab sample directly eluted in the assay buffer and test is required to assess the sensitivity of the assay as a POCT.

The overall sensitivity found in the analytical validation was probably lower than the overall clinical sensitivity because we used fresh samples in the clinical study while the samples for the analytical study were stored during 3 to 6 days at -20°C, and transported at room temperature after thawing in the fridge. As tested with four clinical samples (stored at 2-8°C), the intensity of the reaction slightly diminished after 24h. Further sample storage studies have to be conducted in the near future.

According to the manufacturer’s IFU, the nasopharyngeal samples must be tested as soon as possible after collection, which concords with the need for rapid diagnosis in this pandemic situation. In our population, we did not see any difference between nasopharyngeal swabs and nasopharyngeal aspirates. Because recent published data report high viral loads in nasopharyngeal, mid-turbinate, and nares specimen, we can infer that the ICT could also have a good sensitivity for such specimen. This has previously been described for other viruses and quantitative techniques (20). The evaluation on other swabs such as foam or polyester swabs should also be performed.

When we reflect on the cross-reactivity study, the number of samples analysed for *S. aureus* was not sufficient and sputum was not prescribed as sample type in the manufacturer’s IFU. Whether the false positive sputum was due to a matrix effect, its high-viscosity or to *S. aureus*, is not clear. Sputa from cystic fibrosis patients possess special characteristics, like higher viscosity and multi-pathogenic culture. To be prudent, at this stage, sputum should be avoided with the COVID-19 Ag Respi-Strip, because false-positive results cannot be excluded with this sample type. Whether cystic fibrosis patients should be excluded from analysis with the COVID-19 Ag Respi-Strip and directly receive a diagnosis with qRT-PCR, is not clear, but could be a judicious advice, as long as we lack further evidence.

In a subpopulation with high viral shedding (N=88 with Ct<25), probably the most infectious patients, the clinical sensitivity was 73.9% and we noticed during evaluation that the majority of the positive patients contained high viral loads. Indeed, 66.7% of all positive patients in the clinical performance study had a Ct below 25. Shi Y. et al. mention the severe respiratory symptomatic stage is associated with high viral load (4).

In regard to our results, the test is accurate enough (with an overall accuracy of 82.6%) for implementing in the frame of an integrative diagnostic strategy combining both rapid diagnostic test based on SARS-CoV-2 antigen detection with COVID-19 Ag Respi-Strip, molecular POCT and diagnostics on large automated platform, the latter often requiring a TAT of more than 4 hours.

Figure 2 shows the rapid antigen test can have a role to rapidly take measures at the entry of the emergency department in a pandemic context with high prevalence of COVID-19. Thanks to its specificity of 99.5%, antigen test-positive patients could receive immediate care while antigen test-negative patients will need to wait for CT-scan for triage and the result of qRT-PCR for confirmation of SARS-CoV-2. When the peak of the epidemic approaches and prioritization has to be done, to relieve the emergency department in a situation seeking implementation of fast hygienic measures and rapid patient care, it is defensible not to confirm the antigen test-positive patients with qRT-PCR.

In the proposal for a diagnostic decision algorithm, the TAT could be restrained to 15 minutes after sample collection for patients with a high viral load (Fig. 2). The decision algorithm in Figure 2 shows the potential clinical usefulness of the COVID-19 Ag Respi-Strip for patients suspected of SARS-COV-2 infection thanks to its high positive predictive value of 98.7%. However, suspected patients suffering from severe comorbidities could benefit from molecular POCT if available, because a higher negative predictive value is desired. But the format and the cost of molecular POCT limit them in large screening strategy especially when resources are limited and distribution is delayed.

Since the Hospital Urgency Plan has been declared in Belgium, the healthcare workers focus on severely and critically-ill COVID-19 patients needing hospitalisation. Nasopharyngeal samples for COVID-19 suspected patients are collected at presentation at the emergency department. These samples possess probably the highest viral load because they are the closest to the date of onset of symptoms referring to the kinetics of the viral load as shown in Cao et al. (21).

For this reason, we consider the COVID-19 Ag Respi-Strip could have a relevant place in the diagnostic algorithm at the entry of the emergency department, for the COVID-19 suspected patients at risk of developing severe disease who will require hospitalisation, according to the definition of a possible COVID-19 case as described by the Authorities (Fig. 2).

This decision algorithm would prepare all clinical laboratories for the significant increase in the number of specimen that will need to be tested for COVID-19, when large areas of the country face with community transmission. If a 24/7/365 work organization is proposed by the lab, the rapid result for positive cases in 30 minutes after reception of the specimen, would help to take immediate measures to prevent further spread of the most infectious cases. In a multi-site consolidated clinical microbiology laboratory model such as in the LHUB-ULB (22), a 24-h COVID-19 diagnostic service is provided by dedicated clinical microbiology technologists located in the central lab, while in satellite labs the COVID-19 Ag Respi-Strip is handled 24 hours a day by technologists from chemistry or haematology after cross-training to competently perform and interpret the results of the rapid test. Samples with a negative ICT result are transferred to the central laboratory for molecular diagnosis.

Post-implementation analysis of samples collected in the LHUB-ULB between the 31^st^ of March and the 7^th^ of April 2020 from patients in 4 hospitals from Brussels showed 33.3% (325/975) total positive COVID-19 test results of which 39.7% (129/325) was detected by COVID-19 Ag Respi-Strip. At epidemic peak days with screening of only COVID-19 suspected patients, the proposed algorithm allowed us to avoid 13.2% (129/975) screening qRT-PCR tests, reducing not only expensive qRT-PCR costs, but also the consumption of scarce reagents and consumables. Even if the cost of COVID-19 Ag Respi-Strip is far lower than the cost of our molecular diagnostic methods, budget impact should be studied within a larger health economic frame questioning whether a CT-scan would still be needed for triage and taking into account that the time gain for implementation of biosafety measures when the early positive result of the COVID-19 Ag Respi-Strip will help the clinician at the entry of the emergency to redirect his patient faster without the need for performing a CT-scan.

At this point, no outpatient population has been sampled due to the lack of material for diagnostic testing. Further studies are needed to test the COVID-19 Ag Respi-Strip in the outpatient population, with a special attention to healthcare workers at the frontline like general practitioners (GPs) and pharmacists, in regular contact with mild and pauci-symptomatic patients. One could consider that in conjunction with public health authorities, isolation measures could eventually be focused on those outpatient people with a positive COVID-19 Ag Respi-Strip result, in a subpopulation with a high pre-test prevalence, supposing it could become a tool for managing the lock-down. The implementation of this measure would depend on the precipitating pandemic situation and the possibility to start supplemental prospective outpatient studies or not, regarding the priority in which patient population the COVID-19 Ag Respi-Strip would be implemented first, considering the production capacities of this new method and the highest public health impact of reducing transmission using available resources. Furthermore, the COVID-19 Ag Respi-Strip could be used in residential care centers, supposing a high positivity ratio in closed communities.

Considering risk management and the bottlenecks mentioned in Fig. 3, the Ishikawa diagram shows that COVID-19 Ag Respi-Strip is readily implementable in all clinical laboratories, also the peripheral labs, procuring first-line diagnostic results inside and outside the hospital, to GPs and medical specialists all over the country. Although the standard operating procedure seems to be written for using the device as a POCT, caution has to be taken before widely application in GPs’ cabinets for three reasons. First, in Belgium the legal framework has not been published yet for designating responsibilities of healthcare workers using POCT outside the hospital environment. A proposal for a legal framework has been drafted, closely relating the GP to a clinical laboratory to guarantee traceability within a recognized quality management system and certification of competent users after training. Secondly, regarding the biosecurity rules for handling COVID-19 suspicious respiratory samples, and the absence of a Biological Safety Cabinet Class II at the GP’s office, the GP should possess all protection material like a biohazard bin, sterile gloves, a disposable lab coat, lab glasses and a FFP2 mask. Third, in this study, we did not record how many COVID-19 Ag Respi-Strips gave weak intensities for the control and test lines. But in our User friendly study, the observers mentioned a source of error when reading weakly positive COVID-19 Ag Respi-Strip due to the difficulty to visualize the colour line through the closed tube. To better visualize the strip when good light (such as with Crystal tubes) is not available, the lab technician had sometimes to open the test tube in the laminar air flow cabinet and pull out the strip with forceps. This type of operation would never be acceptable in a GP environment. At this point, the design of the device is a strip into a closed reaction tube, but R&D will continue to improve its maturity, with the perspective of a closed cassette including closed sample handler enhancing visualisation of the test line and reducing the risk of user error in interpretation of the results. Then, a pilot study should evaluate the applicability of the COVID-19 Ag Respi-Strip as a POCT device at the GP’s office, to report the performances obtained in a relevant environment with the intended users.

## CONCLUSION

Our study is the first report evaluating the diagnostic efficacy of the disposable rapid antigen COVID-19 Ag Respi-Strip to detect SARS-COV-2 which expansively spreads worldwide. The satisfying performance and operational utility are promising for first-line diagnosis of COVID-19 in this pandemic situation, accelerating the care process and limiting qRT-PCR analysis to those patients found negative with the rapid antigen test. As mentioned in the proposed diagnostic algorithm, the COVID-19 Ag Respi-Strip may play a triage role during the peak of the pandemic because of the high prevalence, especially at the entry of the emergency department when patients present with high viral load. It is readily implementable in all clinical laboratories, procuring first-line diagnostic results inside and outside the hospital, to GPs and medical specialists. On the contrary, the applicability of the IVD medical device as POCT for GPs could be questioned regarding its maturity (not yet available as a closed cassette with closed sample handler), the legal responsibilities and the concordant distribution of biosafety material, especially when facing worldwide ruptures of FFP2 masks. In summary, the role of the COVID-19 Ag Respi-Strip is complementary to the currently-used molecular techniques.

## Data Availability

Data are available

## ACKNOWLEDGMENTS

We wish to thank the personnel of the LHUB-ULB and Coris BioConcept for its daily technical assistance. UZ Leuven, as national reference centre for respiratory pathogens, is supported by Sciensano, which is gratefully acknowledged. The authors would like to thank Patrick Murray and Alex van Belkum for their generous and always positive support. We would like to acknowledge Viviane Van Hoof, President of the POCT working group at Sciensano. This work is dedicated to the healthcare workers, the patients and families affected by SARS-CoV-2.

